# Incidence and risk factors for post-stroke delirium in the elderly: a national inpatient sample (NIS) analysis

**DOI:** 10.1101/2025.08.13.25333646

**Authors:** Jianrong Zhang, Mei Chen, Yaoyang Huo, Yu’e Wu, Xilin Liu, Xiaohuan Li, Jingqin Wang, Fengling Yang, Gang Liu, Hao Xie, Ying Gao

## Abstract

**Background:** Post-stroke delirium (PSD) is a critical neuropsychiatric condition affecting up to 50% of elderly patients during hospitalization, often leading to poorer outcomes. Despite its prevalence, PSD remains underrecognized in clinical practice, and national-level studies exploring its risk factors are limited.

**Objective:** This study aimed to examine the incidence and risk factors associated with PSD in elderly individuals (≥65 years) using a large, nationally representative dataset.

**Methods:** Data from the Healthcare Cost and Utilization Project National Inpatient Sample (2010–2019) were analyzed. Elderly patients with a primary diagnosis of stroke were selected, and PSD was defined using the International Classification of Diseases, Ninth Revision, Clinical Modification (ICD-9-CM) and ICD-10-CM codes. To determine independent risk factors for PSD, multivariate logistic regression was applied, adjusting for demographic, clinical, and hospital-related variables.

**Results:** Among 1,644,773 elderly stroke patients, the incidence of PSD was 19.5%. Patients with PSD were significantly older, with a median age of 79 years, compared to 78 years in those without PSD (*p* < 0.001). They also experienced prolonged hospital stays (5 days vs. 4 days, *p* < 0.001), incurred greater hospitalization costs ($44,863 vs. $35,787, *p* < 0.001), and exhibited a higher risk of in-hospital mortality (12.6% vs. 7.0%, *p* < 0.001). Significant independent risk factors for PSD included advanced age (≥80 years, odds ratio [OR] = 1.237), three or more comorbidities (OR = 2.049), Black race (OR = 1.113), Asian/Pacific Islander race (OR = 1.060), fluid/electrolyte disorders (OR = 1.902), psychoses (OR = 1.765), sepsis (OR = 2.364), and dysphagia (OR = 1.315).

**Conclusions:** PSD is frequently observed in elderly stroke patients and is associated with adverse clinical outcomes. Advanced age, comorbidities, and stroke-related complications are significant risk factors. These results underscore the importance of developing focused prevention and intervention strategies to enhance outcomes for this high-risk population.

## Introduction

Delirium is a severe acute neuropsychiatric syndrome [1,2], representing a loss of brain function in response to pathophysiological stress and occurring commonly following acute stroke [3]. Previous studies have documented that the prevalence of post-stroke delirium varies widely, ranging from 2% to 66% [4–7]. In addition, a systematic review indicated a clear trend where older age correlates with an increased risk of developing delirium post-stroke, with prevalence rates of 20% for ages 60-64, 25% for 65-74, and 34% for those aged 75-79 [8]. Post-stroke delirium manifests as sudden cognitive disruptions that impact attention, memory, language, and visuospatial processing [9]. Specifically, patients exhibit a reduced capacity to concentrate, sustain, or shift their attention [10]. Post-stroke delirium is closely linked to adverse outcomes. Studies have shown that post-stroke delirium is associated with higher in-hospital mortality (12.3% vs. 7.8%) and a substantial medical cost burden, contributing to healthcare costs exceeding $164 billion annually [3,11]. Furthermore, it is associated with prolonged hospital stays, worsened functional and cognitive impairment, greater rehabilitation needs, and a higher likelihood of transfer to inpatient care [12,13,14]. Consequently, post-stroke delirium poses a significant challenge in stroke management and rehabilitation.

The occurrence of post-stroke delirium is influenced by a complex interplay of risk factors, which can be classified into susceptibility and predisposing factors. Susceptibility factors include advanced age, male gender, pre-stroke cognitive impairment, and various systemic or metabolic diseases [15]. Predisposing factors reported in the literature include a high comorbidity burden, depression, and a history of alcohol abuse [16]. Despite the high susceptibility of stroke patients to delirium, current clinical guidelines often fail to prioritize delirium management in stroke care [17]. This oversight results in inadequate detection and management strategies, emphasizing the need for heightened awareness among healthcare professionals [18]. Addressing post-stroke delirium is therefore crucial [19].

Prior studies investigating post-stroke delirium have predominantly been single-center studies, often focusing on specific geographic regions or patient subgroups, creating a gap in understanding national patterns and demographic variations in the risk of post-stroke delirium [9,20]. This study sought to address this gap by utilizing a large, nationally representative database to explore the incidence and risk factors for delirium in older adults with stroke. The findings from this study are intended to attract widespread attention from clinical experts and provide a robust reference for the prospective management of delirium in this population.

## Material and Methods

### Data Source

Data for this study were obtained from the Healthcare Cost and Utilization Project (HCUP) National Inpatient Sample (NIS), the largest publicly available all-payer inpatient healthcare database in the United States. Funded by the Agency for Healthcare Research and Quality (AHRQ), the NIS employs a stratified sample drawn from over 1,000 hospitals across 46 states, representing approximately 20% of all U.S. hospital admissions each year [49]. The database contains comprehensive information, including patient demographics, admission status, diagnoses, procedures, comorbidities, hospital characteristics, insurance type, length of stay (LOS), in-hospital mortality, total charges, and discharge disposition. All classifications were encoded according to the International Classification of Diseases, Ninth Revision, Clinical Modification (ICD-9-CM) and ICD-10-CM standards. As this study utilized anonymized, publicly available data, it was not subject to ethical approval protocols.

### Data Collection

Elderly patients (aged ≥65 years) hospitalized between January 1, 2010, and December 31, 2019, with a primary diagnosis of stroke (identified using ICD-9-CM and ICD-10-CM codes) were included in this retrospective cohort study. Post-stroke delirium was identified using ICD-9-CM and ICD-10-CM diagnostic codes [see S1 File]. Patients with incomplete data, those under 65 years of age, and those admitted for non-stroke diagnoses were excluded. The final study cohort was stratified into two groups based on the presence or absence of delirium following stroke. Demographic data (age, race, sex), clinical characteristics (length of stay [LOS], hospitalization costs, admission mode), medical complications (e.g., pneumonia, sepsis, acute renal failure, urinary tract infections, acute myocardial infarction), and pre-existing comorbidities (e.g., fluid and electrolyte disorders, psychoses) were collected (Table 1). Fig 1 delineates both the exclusion criteria and the data collection process.

**Fig 1.**
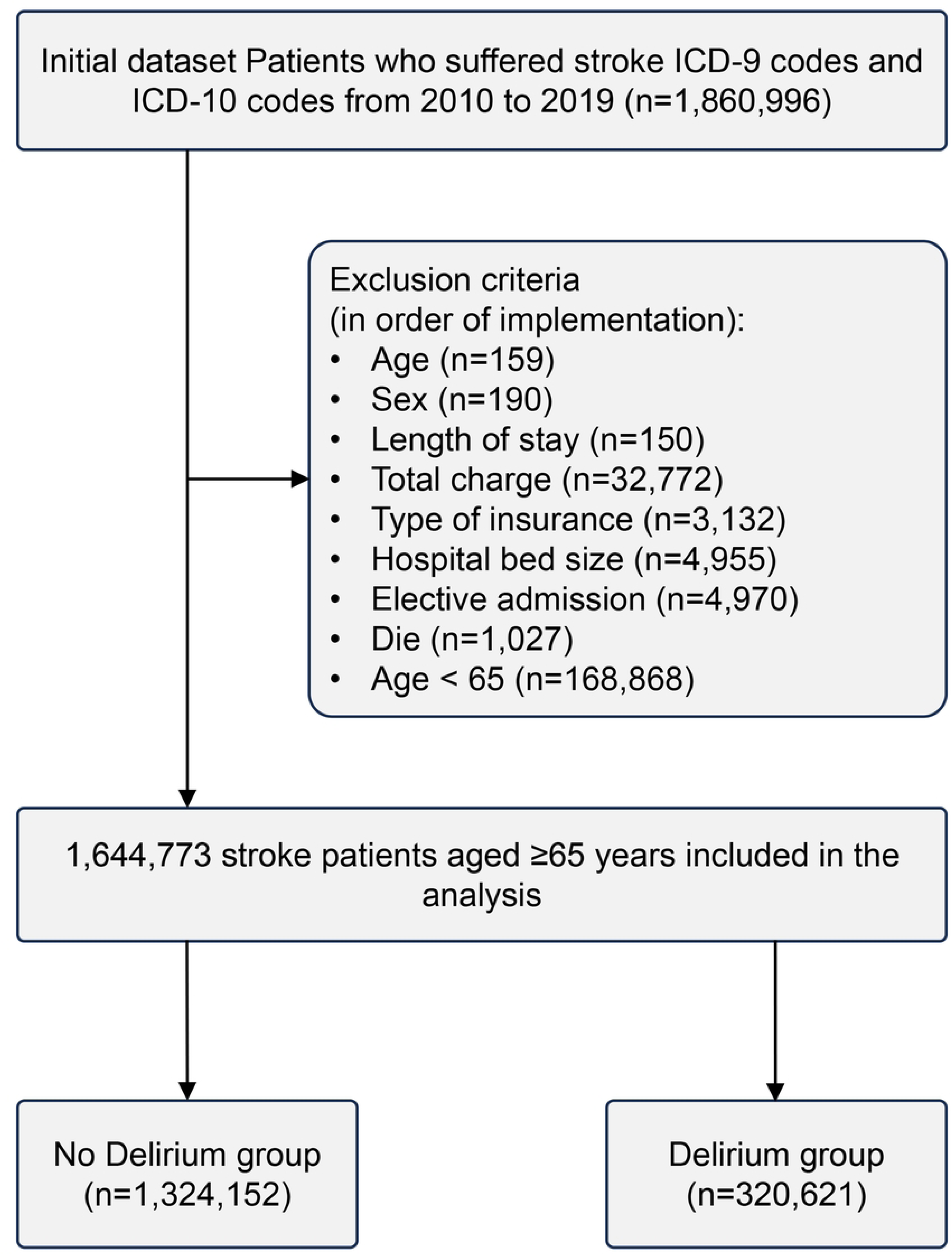
Flowchart depicting the study population selection process.

**Table 1.**
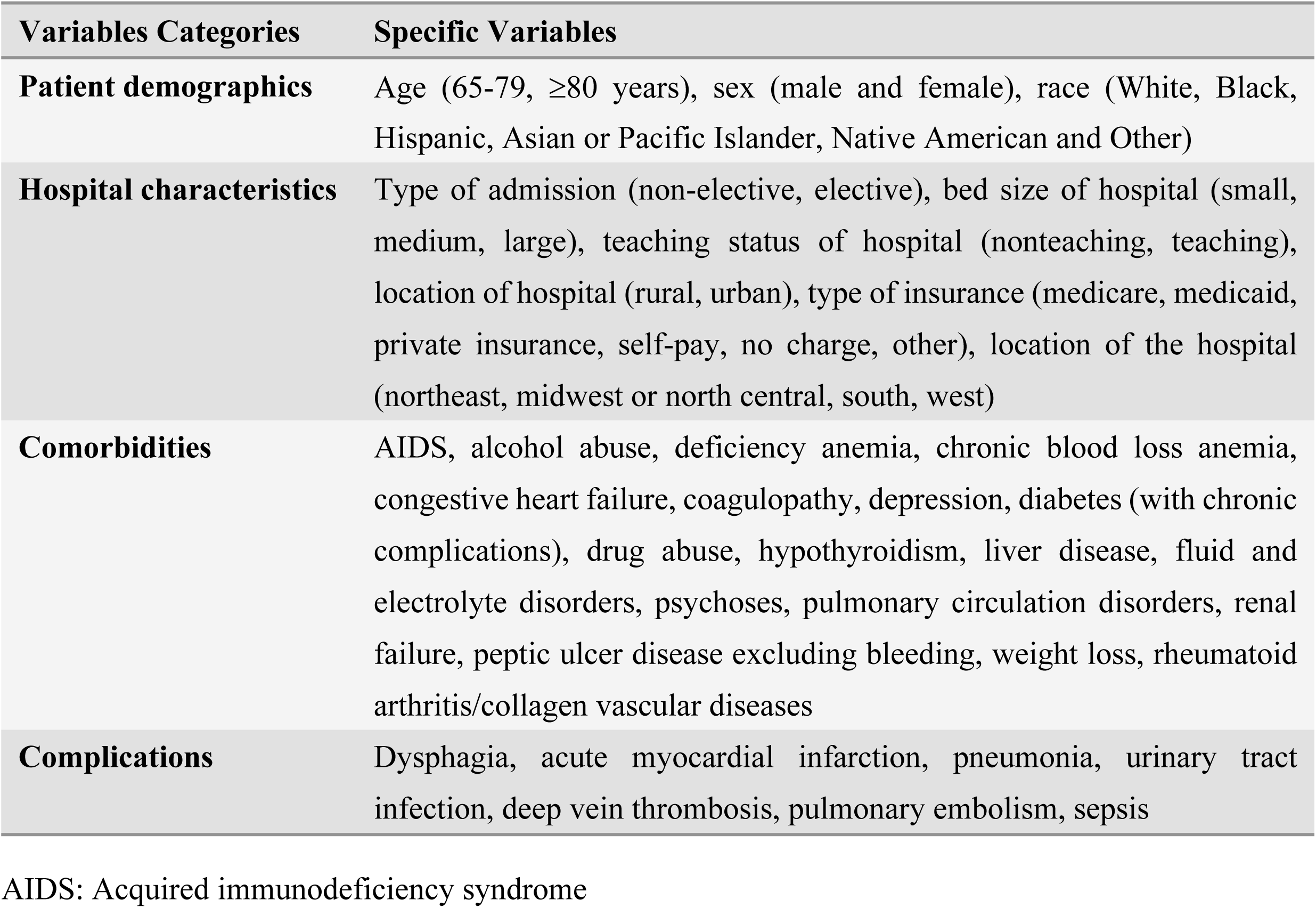
Variables entered into the binary logistic regression analysis.

### Data Analysis

Statistical analyses were performed with SPSS version 25 (IBM SPSS Statistics, USA). The normality of continuous variables was evaluated using the Kolmogorov-Smirnov test. We applied non-parametric Wilcoxon rank-sum tests for continuous variables and chi-square or Fisher’s exact tests for categorical variables. To identify independent risk factors for delirium following stroke, both univariate and multivariate logistic regression analyses were conducted. These analyses adjusted for demographic factors, comorbidities, hospital characteristics, and complications. Odds ratios (OR) with 95% confidence intervals (CI) were computed. Considering the extensive sample sizes in previous NIS studies, statistical significance was defined as *p≤*0.001 [49].

## Results

### Characteristics related to demographics and hospital factors for patients in both groups

Between 2010 and 2019, the NIS database recorded 1,644,773 elderly stroke patients. Of these, 320,621 cases of PSD were documented, corresponding to an incidence rate of 19.5% (Table 2). Moreover, patients who developed delirium were significantly older, with a median age of 79 years, compared to 78 years among those without delirium (*p* < 0.001). The delirium group exhibited a higher proportion of patients aged 80 years and above (43.5% vs. 43.2%, *p* < 0.001). A significant difference in gender distribution was observed between the two groups (*p* = 0.001). Regarding race, a higher proportion of Black patients (11.7% vs. 10.4%, *p* < 0.001) and Asian/Pacific Islander patients (2.7% vs. 2.5%, *p* < 0.001) developed delirium compared to the non-delirium group. Patients in the delirium group exhibited a significantly extended median hospital stay (5 days vs. 4 days, *p* < 0.001) and incurred greater total hospital costs ($44,863 vs. $35,787, *p* < 0.001). Furthermore, patients with delirium experienced a higher in-hospital mortality rate (12.6% vs. 7.0%, *p* < 0.001) (Table 2) (Fig 2) (Fig 3).

**Fig 2.**
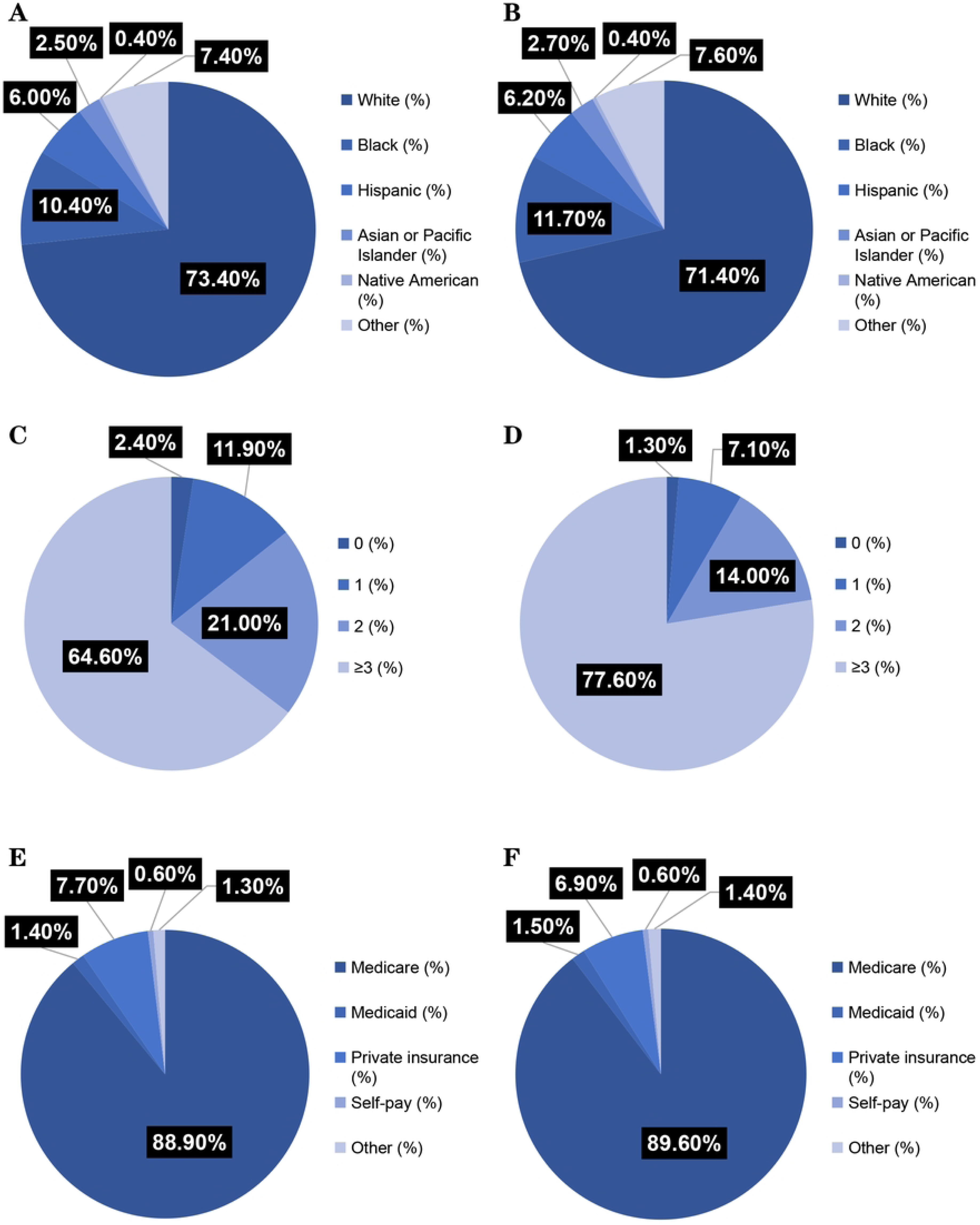
Comparison of patient demographics and hospital characteristics between the two groups. **A** Racial distribution analysis among patients without delirium. **B** Racial distribution analysis among those with delirium. **C** Evaluation of comorbidity burden in non-delirium patients. **D** Assessment of comorbid conditions in patients experiencing delirium. **E** Analysis of insurance types for non-delirium patients. **F** Analysis of insurance types for patients with delirium.

**Fig 3.**
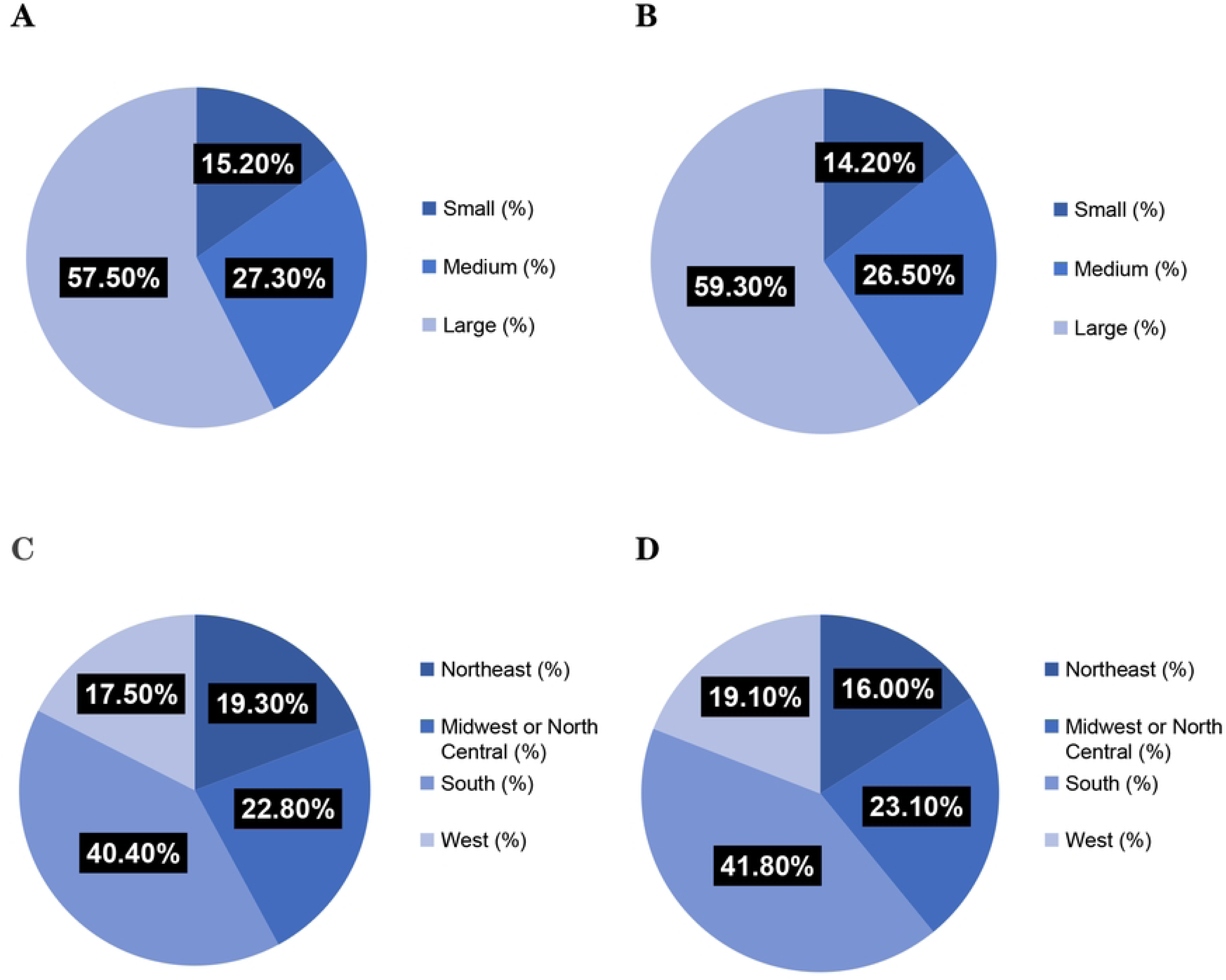
Patient demographics and hospital characteristics between the two groups. **A** Analysis of the number of hospital bed size for non-delirium patients. **B** Analysis of the number of hospital bed size for patients with delirium. **C** Regional distribution analysis for hospitals housing non-delirium patients. **D** Regional distribution analysis for hospitals housing patients with delirium.

**Table 2.** Patient characteristics and outcomes of post-stroke delirium (2010–2019)

| Characteristics | No delirium | delirium | *p* |
| --- | --- | --- | --- |
| **Total (n=count)** | 1,324,152 | 320,621 |  |
| **Total incidence (%)** | 80.5 | 19.5 |  |
| **Age (median, years)** | 78.0 (71.0-84.0) | 79.0 (72.0-86.0) | <0.001 |
| **Age group (%)** |  |  |  |
| 65-79 | 56.80 | 51.50 | <0.001 |
| ≥80 | 43.20 | 43.50 |  |
| **Sex (%)** |  |  |  |
| Male | 46.90 | 45.40 | <0.001 |
| Female | 53.10 | 54.60 |  |
| **Race (%)** |  |  |  |
| White | 73.40 | 71.40 | <0.001 |
| Black | 10.40 | 11.70 |  |
| Hispanic | 6.00 | 6.20 |  |
| Asian or Pacific Islander | 2.50 | 2.70 |  |
| Native American | 0.40 | 0.40 |  |
| Other | 7.40 | 7.60 |  |
| Number of Comorbidities (%) |  |  |  |
| 0 | 2.40 | 1.30 |  |
| 1 | 11.90 | 7.10 | <0.001 |
| 2 | 21.00 | 14.00 |  |
| ≥3 | 64.60 | 77.60 |  |
| LOS (median, d) | 4 (2-6) | 5 (3-10) | <0.001 |
| TOTCHG (median, \$) | 35,787<br>(20,633-67,170) | 44,863<br>(23,224-95,960) | <0.001 |
| Type of insurance (%) |  |  |  |
| Medicare | 88.90 | 89.60 |  |
| Medicaid | 1.40 | 1.50 |  |
| Private insurance | 7.70 | 6.90 | <0.001 |
| Self-pay | 0.60 | 0.60 |  |
| No charge | 0.00 | 0.00 |  |
| Other | 1.30 | 1.40 |  |
| Bed size of hospital (%) |  |  |  |
| Small | 15.20 | 14.20 |  |
| Medium | 27.30 | 26.50 | <0.001 |
| Large | 57.50 | 59.30 |  |
| Elective admission (%) | 87.10 | 91.60 | <0.001 |
| Type of hospital (teaching %) | 61.10 | 61.30 | 0.041 |
| Location of hospital (urban, %) | 90.50 | 90.90 | <0.001 |
| Region of hospital (%) |  |  |  |
| Northeast | 19.30 | 16.00 |  |
| Midwest or North Central | 22.80 | 23.10 | <0.001 |
| South | 40.40 | 41.80 |  |
| West | 17.50 | 19.10 |  |
| Died (%) | 7.00 | 12.60 | <0.001 |

LOS: Length of stay, TOTCHE: Total charge

### Post-Stroke Complications and Delirium

Patients with post-stroke delirium exhibited significantly higher rates of various medical complications, including dysphagia (16.9%), acute myocardial infarction (6.9%), pneumonia (13.0%), urinary tract infection (21.8%), deep vein thrombosis (2.5%), pulmonary embolism (1.3%), and sepsis (11.5%), compared to those without post-stroke delirium (*p* < 0.001) (Table 4). Furthermore, multivariate logistic regression analysis indicated an association between post-stroke delirium and dysphagia (OR=1.315; 95% confidence interval [CI] = 1.301–1.329), acute myocardial infarction (OR=1.157; 95% CI = 1.139–1.176), pneumonia (OR = 1.434; 95% CI = 1.416–1.453), urinary tract infection (OR = 1.685; 95% CI = 1.668–1.702), deep vein thrombosis (OR = 1.270; 95% CI = 1.235–1.307), pulmonary embolism (OR = 1.132; 95% CI = 1.090–1.176), and sepsis (OR = 2.364; 95% CI = 2.329–2.400) (Table 5).

**Table 3.**
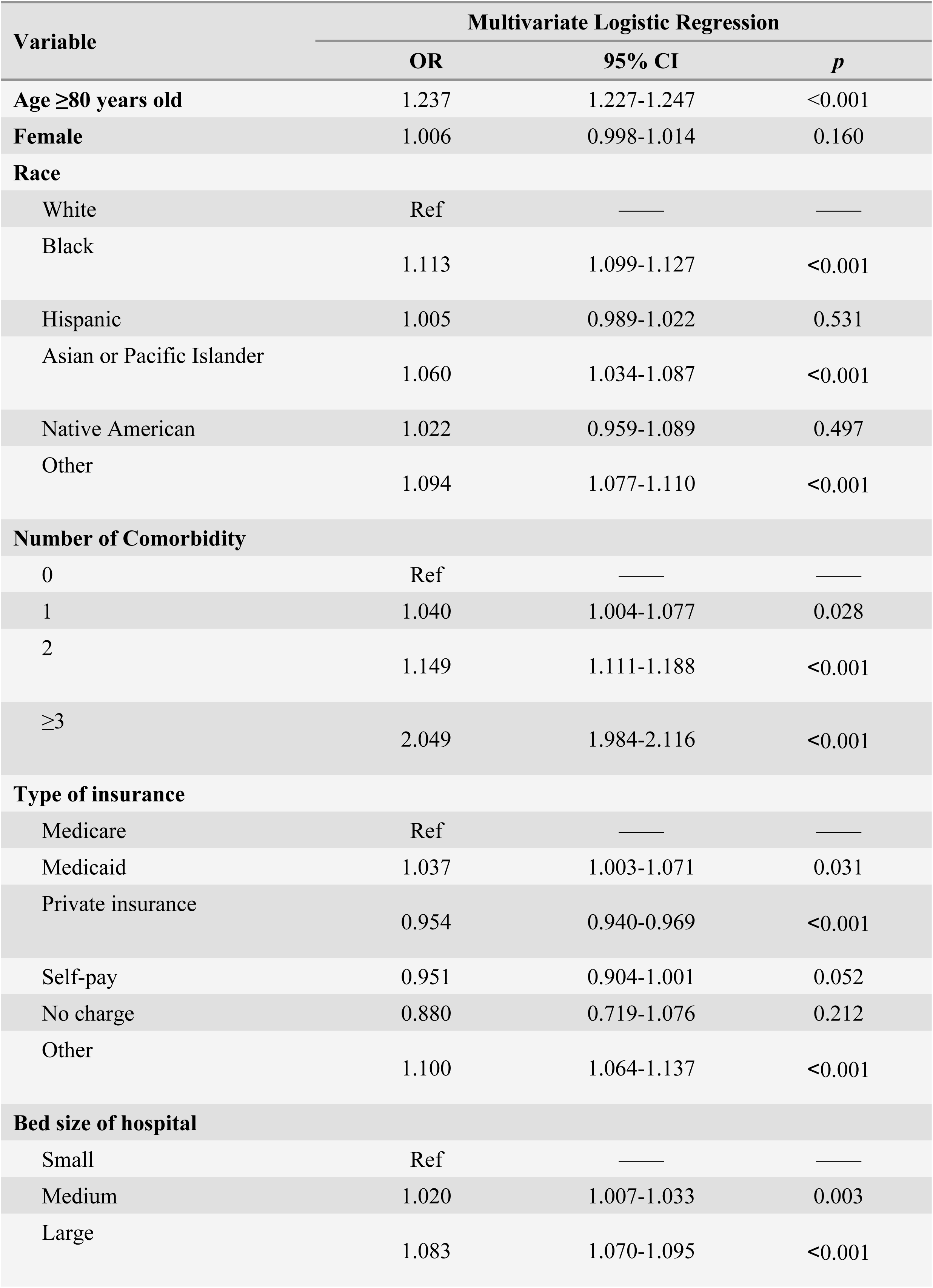
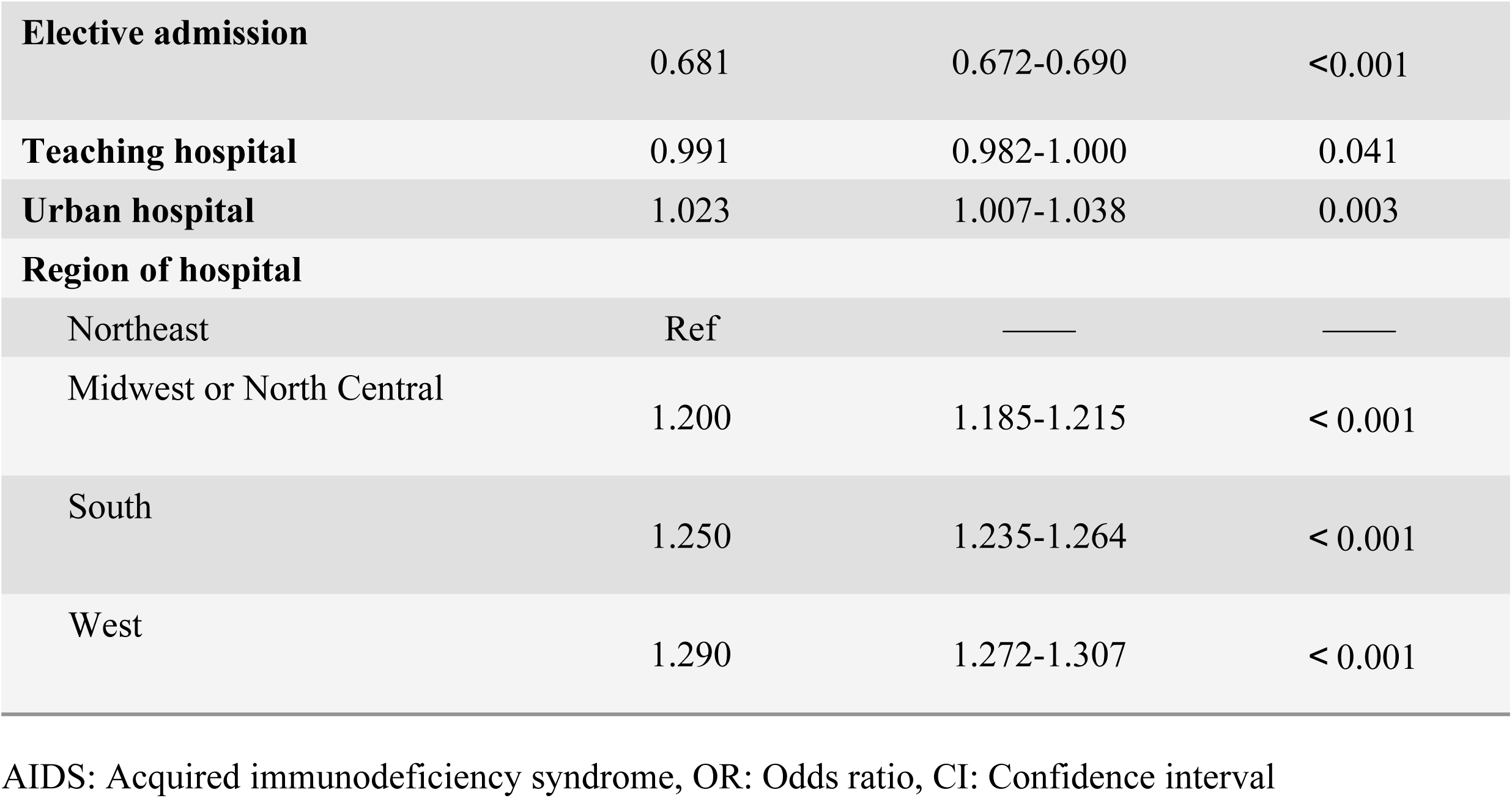
Risk factors associated with post-stroke delirium.

**Table 4.** Relationship between pre-stroke comorbidities and post-stroke delirium.

| Comorbidities | Univariate Analysis |  |  | Multivariate Logistic Regression |  |  |
| --- | --- | --- | --- | --- | --- | --- |
|  | No delirium | Delirium | *p* | OR | 95% CI | *p* |
| **Prestroke comorbidities, n (%)** |  |  |  |  |  |  |
| **Acquired immune deficiency syndrome** | 1,264 (0.1) | 308 (0.1) | 0.921 | 0.851 | 0.749-0.966 | 0.013 |
| **Alcohol abuse** | 32,064 (2.4) | 9,802 (3.10) | < 0.001 | 1.126 | 1.099-1.153 | < 0.001 |
| **Deficiency anemia** | 132,420 (10.0) | 42,062 (13.1) | < 0.001 | 1.150 | 1.136-1.164 | < 0.001 |
| **Rheumatoid arthritis/collagen vascular diseases** | 34,500 (2.60) | 8,817 (2.7) | < 0.001 | 1.023 | 0.999-1.048 | 0.063 |
| **Chronic blood loss anemia** | 8,876 (0.7) | 2,435 (0.8) | < 0.001 | 0.956 | 0.913-1.001 | 0.056 |
| **Congestive heart failure** | 227,917 (17.2) | 69,967 (21.8) | < 0.001 | 1.186 | 1.174-1.198 | < 0.001 |
| **Coagulopathy** | 74,568 (5.6) | 28,720 (9.0) | < 0.001 | 1.333 | 1.314-1.353 | < 0.001 |
| **Depression** | 132,876 (10.0) | 39,377 (12.3) | < 0.001 | 1.236 | 1.221-1.251 | < 0.001 |
| **Diabetes with chronic complications** | 169,315 (12.8) | 47,067 (14.7) | < 0.001 | 1.062 | 1.050-1.075 | < 0.001 |
| **Drug abuse** | 7,988 (0.6) | 2,965 (0.9) | < 0.001 | 1.340 | 1.283-1.400 | < 0.001 |
| **Hypothyroidism** | 225,079 (17.0) | 56,172 (17.5) | < 0.001 | 1.012 | 1.002-1.023 | 0.019 |
| **Liver disease** | 21,184 (1.6) | 6,757 (2.1) | < 0.001 | 1.036 | 1.007-1.066 | 0.015 |
| **Fluid and electrolyte disorders** | 306,051 (23.1) | 124,579 (38.9) | < 0.001 | 1.902 | 1.886-1.918 | < 0.001 |
| **Psychoses** | 25,367 (1.9) | 11,164 (3.5) | < 0.001 | 1.765 | 1.725-1.806 | < 0.001 |
| **Pulmonary circulation disorders** | 52,698 (4.0) | 15,224 (4.7) | < 0.001 | 1.008 | 0.989-1.027 | 0.439 |
| **Renal failure** | 257,135 (19.4) | 72,822 (22.7) | < 0.001 | 1.059 | 1.048-1.070 | < 0.001 |
| **Peptic ulcer disease excluding bleeding** | 4,398 (0.3) | 1,370 (0.4) | < 0.001 | 1.092 | 1.026-1.162 | 0.006 |
| **Weight loss** | 70,018 (5.3) | 34,322 (10.7) | < 0.001 | 1.690 | 1.667-1.714 | < 0.001 |

OR: Odds ratio, CI: Confidence interval

**Table 5.**
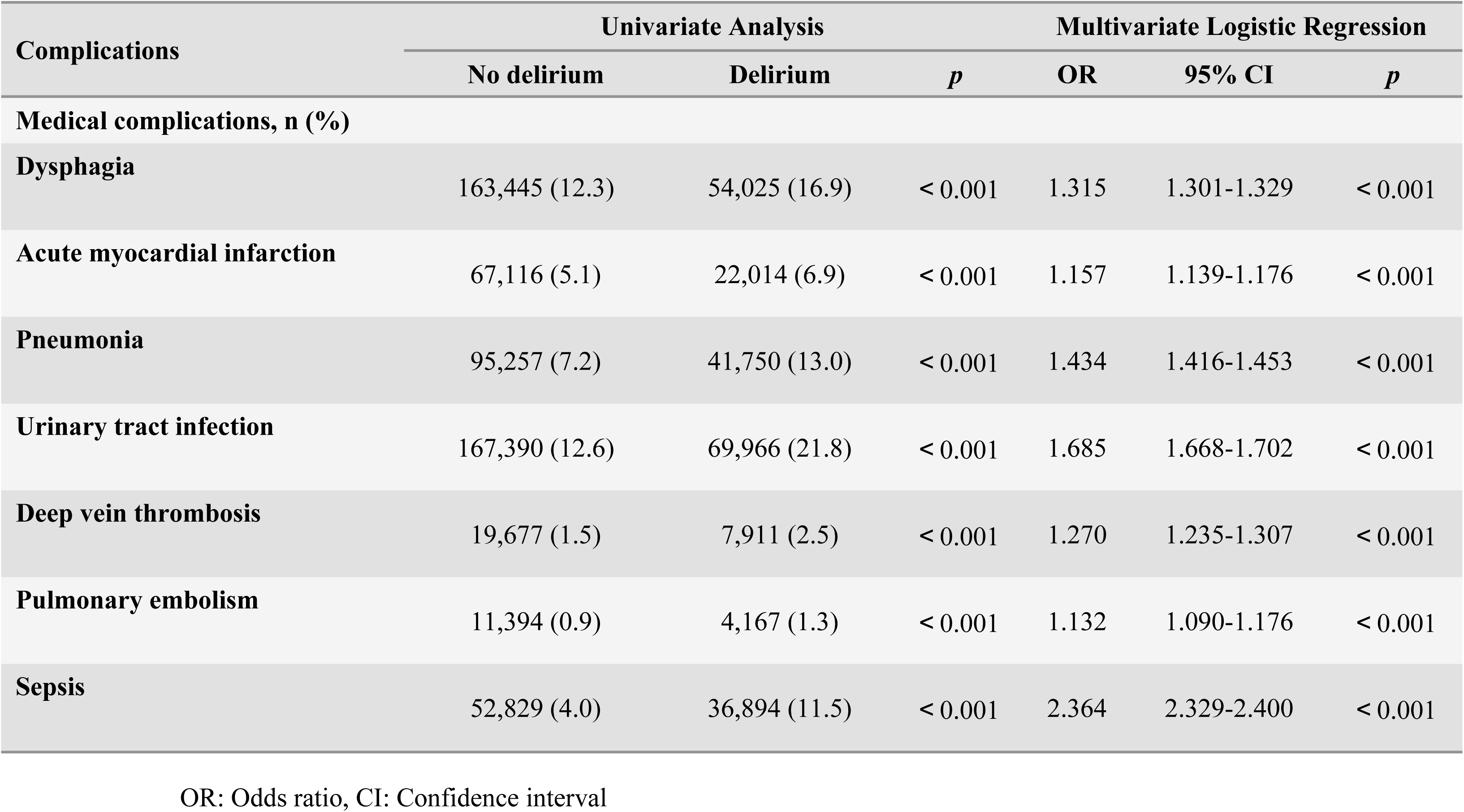
Relationship between post-stroke delirium and post-stroke complications.

| Complications | Univariate Analysis |  |  | Multivariate Logistic Regression |  |  |
| --- | --- | --- | --- | --- | --- | --- |
|  | No delirium | Delirium | *p* | OR | 95% CI | *p* |
| **Medical complications, n (%)** |  |  |  |  |  |  |
| **Dysphagia** | 163,445 (12.3) | 54,025 (16.9) | < 0.001 | 1.315 | 1.301-1.329 | < 0.001 |
| **Acute myocardial infarction** | 67,116 (5.1) | 22,014 (6.9) | < 0.001 | 1.157 | 1.139-1.176 | < 0.001 |
| **Pneumonia** | 95,257 (7.2) | 41,750 (13.0) | < 0.001 | 1.434 | 1.416-1.453 | < 0.001 |
| **Urinary tract infection** | 167,390 (12.6) | 69,966 (21.8) | < 0.001 | 1.685 | 1.668-1.702 | < 0.001 |
| **Deep vein thrombosis** | 19,677 (1.5) | 7,911 (2.5) | < 0.001 | 1.270 | 1.235-1.307 | < 0.001 |
| **Pulmonary embolism** | 11,394 (0.9) | 4,167 (1.3) | < 0.001 | 1.132 | 1.090-1.176 | < 0.001 |
| **Sepsis** | 52,829 (4.0) | 36,894 (11.5) | < 0.001 | 2.364 | 2.329-2.400 | < 0.001 |

OR: Odds ratio, CI: Confidence interval

### Risk factors associated with delirium after stroke in the elderly

Multivariate logistic regression analysis identified several significant independent risk factors for post-stroke delirium (Table 3 and Table 4): age ≥80 years (OR=1.237; 95% CI== 1.227–1.247, *p* < 0.001), Black race (OR = 1.113; 95% CI = 1.099–1.127), Asian or Pacific Islander race (OR = 1.060; 95% CI = 1.034–1.087), Other race (OR = 1.094; 95% CI = 1.077–1.110), ≥3 comorbidities (OR = 2.049; 95% CI = 1.984–2.116), large hospital bed size (OR = 1.083; 95% CI = 1.070–1.095), elective admission (OR = 0.681; 95% CI = 0.672–0.690), geographic region: Midwest/North Central (OR = 1.200; 95% CI = 1.185–1.215), South (OR = 1.250; 95% CI = 1.235–1.264), and West (OR = 1.290; 95% CI = 1.272– 1.307), alcohol abuse (OR = 1.126; 95% CI = 1.099–1.153), deficiency anemia (OR = 1.150; 95% CI = 1.136–1.164), congestive heart failure (OR = 1.186; 95% CI = 1.174–1.198), coagulopathy (OR = 1.333; 95% CI = 1.314–1.353), depression (OR = 1.236; 95% CI = 1.221–1.251), diabetes with chronic complications (OR = 1.062; 95% CI = 1.050–1.075), drug abuse (OR = 1.340; 95% CI = 1.283–1.400), fluid and electrolyte disorders (OR = 1.902; 95% CI = 1.886–1.918), psychoses (OR = 1.765; 95% CI = 1.725–1.806), renal failure (OR = 1.059; 95% CI = 1.048–1.070), and weight loss (OR = 1.690; 95% CI = 1.667–1.714).

## Discussion

PSD is a frequently observed neuropsychiatric complication among elderly individuals who have suffered a stroke. It is linked to adverse outcomes, including extended hospital stays, higher mortality rates, and delayed recovery. Factors such as advanced age, existing comorbid conditions, and stroke-related complications significantly elevate the risk of PSD, highlighting the importance of tailored preventive and management approaches. A large cohort of elderly stroke patients was examined to determine the incidence and risk factors associated with PSD. Our study determined an overall incidence of post-stroke delirium of 19.5% (Table 2), lower than the reported prevalence in the literature (30%) [21]. Besides, Doris et al. reported a 27.4% prevalence among stroke patients over 50 years old [22]. Discrepancies between studies may arise from limitations in prior research, such as small sample sizes and an overrepresentation of elderly patients. These factors may have led to an inflated estimation of delirium prevalence following acute stroke. Moreover, variations in diagnostic criteria and the accuracy of delirium identification across researchers and institutions may influence the reliability of reported outcomes.

Our study highlights the significant impact of post-stroke delirium on clinical outcomes. Patients with delirium experienced extended hospital stays (5 days vs. 4 days, *p* < 0.001), increased hospitalization expenses ($44,863 vs. $35,787, *p* < 0.001), and elevated in-hospital mortality rates (12.6% vs. 7.0%, *p* < 0.001) (Table 2). These results aligned with prior research indicating that delirium is a prevalent and severe complication in stroke patients, contributing to cognitive and functional decline, prolonged hospitalization, and increased mortality risk [23,24,25].

Our analysis revealed racial disparities, with Black and Asian/Pacific Islander patients demonstrating an increased risk of developing delirium (Table 3). These findings aligned with previous studies emphasizing racial disparities in health outcomes [26,27]. Factors such as cultural, socioeconomic, and healthcare access differences may contribute to these observed disparities. Our results suggest that demographic and clinical factors should be considered in the management of stroke patients to tailor individualized care strategies [28].

Multiple studies on post-stroke delirium have highlighted the critical role of early detection and intervention in managing risk factors to enhance patient outcomes [29,30]. Consequently, understanding these risk factors is essential for developing targeted interventions. Our findings are largely consistent with previous research and provide further insights into the predictors of post-stroke delirium. Logistic regression analysis indicated that individuals aged 80 years or older (OR = 1.237) had a significantly higher likelihood of developing delirium (Table 3). This finding aligned with prior research emphasizing advanced age as a critical risk factor for delirium [38]. The mechanisms underlying this association are complex and may involve age-related physiological changes in brain function, as well as increased susceptibility to metabolic disturbances during stroke recovery [39,40]. Furthermore, the likelihood of delirium was elevated in patients presenting with three or more comorbidities (OR = 2.05) (Table 3). This finding aligned with previous studies indicating that the presence of multiple comorbid conditions correlates with a poorer stroke prognosis [31,32]. Among these comorbidities, alcohol abuse (OR=1.126) and deficiency anemia (OR=1.150) were associated with delirium (Table 4), indicating that these conditions may exacerbate the neurocognitive decline in stroke patients [33,34,35].

As expected, fluid and electrolyte disorders exhibited the strongest association with post-stroke delirium, with an odds ratio (OR=1.902) (Table 4), corroborating prior evidence that fluid and electrolyte disturbances can affect neuronal function, impair the nervous system, and significantly increase the risk of delirium [36,34,37]. Moreover, a prior history of neuropsychiatric disorders, such as psychoses (OR = 1.765) and depression (OR = 1.236), was associated with an elevated risk of delirium (Table 4). This finding underscores the contribution of pre-existing brain pathology to delirium development [41,34,42]. Systemic comorbidities were also identified as independent risk factors. Renal failure (OR=1.059) and congestive heart failure (OR = 1.186) have been previously linked to delirium (Table 4), likely due to their impact on cerebral perfusion and metabolic stability [43,44,45]. These findings emphasize the necessity of optimizing comorbidity management to mitigate delirium risk and enhance the overall prognosis in elderly stroke patients. Although several risk factors, such as advanced age, prior neurological disorders, and psychiatric comorbidities, are non-modifiable, their identification remains clinically significant. Recognizing high-risk patients allows for targeted delirium prevention strategies, such as enhanced monitoring, early mobilization, and cognitive interventions.

Notably, our study identified private insurance as a protective factor against post-stroke delirium (Table 3). A potential explanation is that, among elderly stroke patients, private insurance often correlates with a favorable financial status, which may facilitate timely access to medical care. Delays in treatment are known to increase the risk of delirium, while adequate financial resources likely contribute to both its prevention and management. Moreover, logistic regression analysis identified elective admission as another protective factor (Table 3), as patients admitted electively tend to be in relatively stable condition. Elective admissions provide patients with the opportunity to prepare in advance, alleviating stress and anxiety upon hospital admission. This psychological stabilization may contribute to a lower incidence of delirium.

The occurrence of delirium was significantly influenced by post-stroke complications. Patients experiencing post-stroke complications such as dysphagia, pneumonia, urinary tract infection, and sepsis exhibited a higher risk of delirium (Table 5) [46,47,48]. The literature has consistently shown that other complications in hospitalized stroke patients tend to induce or worsen delirium [38,48]. Our findings suggest that the management of these complications should be prioritized in the prevention of post-stroke delirium [24].

While this research offers critical insights into the incidence and risk factors associated with post-stroke delirium, several inherent limitations must be acknowledged. The study’s retrospective design restricts our ability to establish causal relationships, and the use of administrative data may have contributed to underreporting or inaccuracies in the coding of variables such as delirium. Moreover, the study did not capture detailed information regarding patient-specific delirium management strategies, which could have influenced the observed outcomes. Additional prospective research is required to validate these results and investigate the mechanisms through which factors like age and comorbidities influence the development of post-stroke delirium.

## Conclusions

This large-scale study of elderly stroke patients (n = 1,644,773) revealed that post-stroke delirium occurred in 19.5% of patients and identified several significant risk factors. Advanced age (≥80 years), increased comorbidity burden (particularly ≥3 comorbidities, OR = 2.049), and specific disorders, such as fluid and electrolyte disorders (OR = 1.902) and psychosis (OR = 1.765), were strongly associated with the development of delirium. Patients with delirium exhibited a higher mortality rate (12.6% vs. 7.0%), longer hospitalization, and an increased incidence of complications, notably sepsis (OR = 2.364). These results highlight the importance of promptly identifying patients at elevated risk and implementing tailored preventive measures within stroke management protocols for the elderly population.

## Abbreviations

PSD: Post-stroke delirium
HCUP: Healthcare Cost and Utilization Project
NIS: National Inpatient Sample
AHRQ: Agency for Healthcare Research and Quality
LOS: Length of stay
AIDS: Acquired immunodeficiency syndrome
OR: Odds ratios
CI: Confidence intervals
ICD-9-CM: International Classifcation of Diseases (Ninth Edition) Clinical Modifcation
ICD-10-CM: International Classifcation of Diseases (Tenth Edition) Clinical Modifcation

## Ethics approval and informed consent

Not applicable. This article does not report any studies involving human subjects or animals. As it exclusively utilizes identified publicly available data, no consent was required, and the Dongguan Houjie Hospital Medical Ethics Committee deemed the study exempt. Therefore, no permission is needed in the Ethics Approval and Consent to Participate section. All methods were performed in accordance with the relevant guidelines and regulations.

## Consent for publication

Not applicable.

## Data availability

This research relies on data from the Nationwide Inpatient Sample (NIS) database, a component of the Healthcare Cost and Utilization Project under the Agency for Healthcare Research and Quality. The NIS is one of the largest publicly available full-payer inpatient care databases in the United States, with documentation available at https://www.hcup-us.ahrq.gov/db/nation/nis/nisdbdocumentation.jsp. Consequently, the authors are unable to share individual or aggregated data.

## Authors’ contributions

JZ, MC, and YH were responsible for the study design, data collection and analysis, results interpretation, and manuscript drafting and revision. YW, XL (Liu), XL (Li), and JW contributed to the study design, results interpretation, and manuscript review. FY and GL handled data collection, data analysis, and also reviewed the manuscript. HX and YG contributed to the study design, results interpretation, and manuscript review. JZ, MC and YH contributed equally to this work, they are both the first authors. HX and YG and are corresponding authors. All authors have read and approved the final manuscript and have agreed on the journal to which it will be submitted.

## Data Availability

The data underlying the results presented in the study are available from https://www.hcup-us.ahrq.gov/db/nation/nis/nisdbdocumentation.jsp.

## Acknowledgements

Not applicable.

---

S1 File

Supplemental Digital Content (Table S1): International Classification of Diseases, 9th Revision, Clinical Modification and Procedure Coding System (ICD-9 CM/PCS) Codes

**That Were Used**.

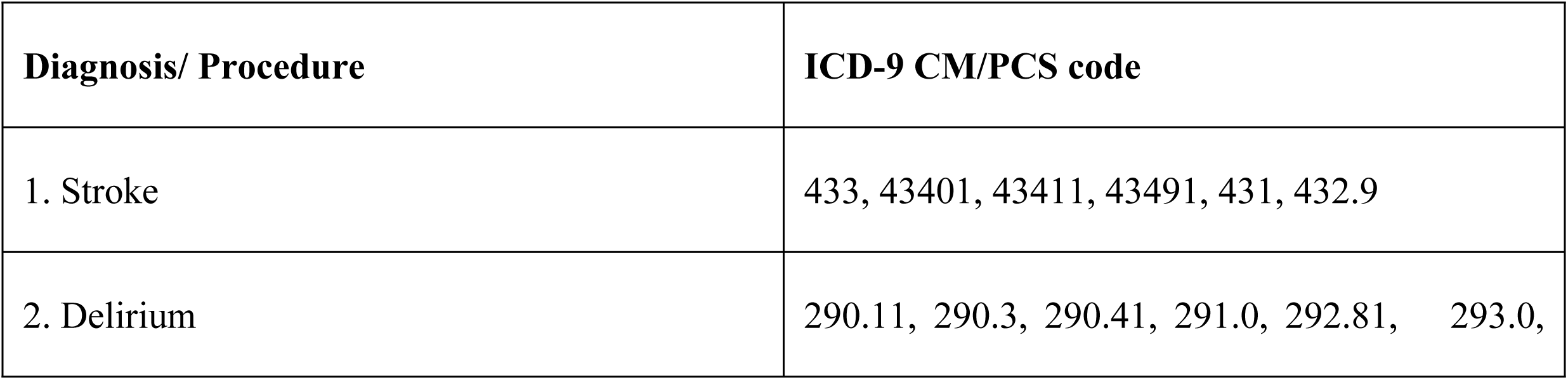
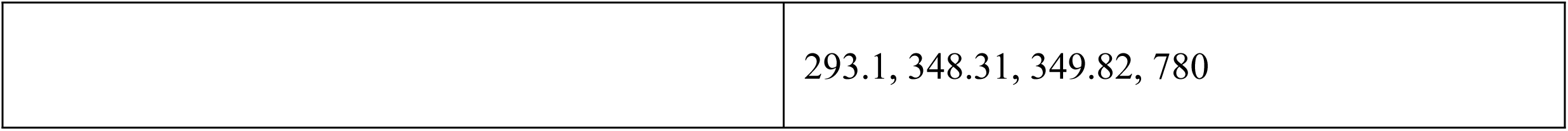

| Diagnosis/ Procedure | ICD-9 CM/PCS code |
| --- | --- |
| 1. Stroke | 433, 43401, 43411, 43491, 431, 432.9 |
| 2. Delirium | 290.11, 290.3, 290.41, 291.0, 292.81, 293.0, |
|  | 293.1, 348.31, 349.82, 780 |

Supplemental Digital Content (Table S2): International Classification of Diseases, 10th Revision, Clinical Modification and Procedure Coding System (ICD-10 CM/PCS) Codes

**That Were Used.**

| **Diagnosis/ Procedure** | **ICD-10 CM/PCS code** |
| --- | --- |
| 1. Stroke | I61, I62.9, I63 |
| 2. Delirium | R41.x*, R41.81, F01.51, F02.81, F03.91, F05.x*,<br>F06.x*, F06.2, F06.8, G93.4, G93.49, G93.41,<br>G92.9, G92.8 |

\*Abbreviations: .x: including subcodes, .00-.99

